# Rapid detection of SARS-CoV-2 antibodies in oral fluids

**DOI:** 10.1101/2020.10.12.20210609

**Authors:** Melanie A. MacMullan, Prithvi Chellamuthu, Aubree Mades, Sudipta Das, Fred Turner, Vladimir I Slepnev, Albina Ibrayeva

## Abstract

Current commercially available methods for reliably detecting antibodies against SARS-CoV-2 remain expensive and inaccessible due to the need for whole blood collection by highly trained phlebotomists using personal protective equipment (PPE). We evaluated an antibody detection approach utilizing the OraSure® Technologies’ Oral Antibody Collection Device (OACD) and their proprietary SARS-CoV-2 total antibody detection enzyme-linked immunosorbent assay (ELISA). We found that the OraSure® test for total antibody detection in oral fluid had comparable sensitivity and specificity to serum-based ELISAs while presenting a more affordable and accessible system with the potential for self-collection.

## Introduction

The COVID-19 pandemic has had a severe impact on populations and economies worldwide (1). While the relationship between antibodies and their protective role against reinfection of SARS-CoV-2 is under investigation, it has been shown that antibodies consistently develop as part of the immune response against the virus in some, if not most, infected individuals (2). Neutralizing antibodies were effective and sustained for at least two and potentially up to twelve years against SARS-CoV, the first SARS coronavirus, strongly suggesting the potential for significant immunity conferred by antibodies against SARS-CoV-2 and prompting extensive studies to identify neutralizing antibodies for therapeutic development (3,4). Further, although residues of the receptor binding domains (RBD) of SARS-CoV and SARS-CoV-2 differ by about 55% (18/33), it has been shown that neutralizing antibodies against SARS-CoV are reactive against SARS-CoV-2. This response indicates overlapping epitopes as the target of these neutralizing antibodies, confirming similarity between antibodies developed against the two SARS-CoVs and supporting the hypothesis of antibody efficacy against re-infection by SARS-CoV-2 (5).

Since the introduction of SARS-CoV-2 in late 2019, individuals infected by the virus have remained immune against re-infection in all but a few sporadic cases. Even in those patients, it is still not clear whether it was a case of re-infection or re-activation of the virus, suggesting that individuals who become infected with SARS-CoV-2 seem to have at least some transient, temporal immunity (6). Given those findings, we have ample reason to believe that the monitoring of antibodies is crucial to understanding global disease spread, immunity against re-infection by the virus, and the effectiveness of potential vaccine candidates.

The industry standard for antibody detection relies on the collection and analysis of patient serum, requiring trained phlebotomists to draw the whole blood from a patient following suspected viral infection (7). That process is time-consuming, costly, and potentially hazardous to the phlebotomist. There is also a substantial need to protect phlebotomists with personal protective equipment (PPEs) such as K95 masks, face shields, gloves etc. that have had limited availability during the pandemic (8). In addition to the health concerns, the requirement of a phlebotomist is a bottleneck for patient blood collection which prevents easy antibody testing for widespread monitoring. Thus, a non-invasive technology for self-collection is needed to improve access to antibody testing.

We have previously demonstrated that patient saliva, instead of serum, can serve as a sufficient medium for the detection of antibodies against SARS-CoV-2 (9). However, given the substantial need for sensitive and specific antibody detection, we did not feel that our method of collection was adequately sensitive for practical manufacturing and mass production for widespread testing. This led us to a collaboration with OraSure® Technologies, who have previously demonstrated the capacity of their Food and Drug Administration-approved OraSure® Oral Antibody Collection Device (OACD) to sample oral fluid antibody against human immunodeficiency virus (HIV) and other pathogens. OraSure® Technologies has adapted the use of this device for better collection of oral specimens and developed an ELISA for detection of antibodies (IgG, IgM, IgA) against SARS-CoV-2 in these specimens (10,11)

## Results

To determine whether we could adequately detect antibodies against SARS-CoV-2 in oral fluid self-collected under observation using the OraSure® OACD (Schematic 1), we evaluated paired serum and oral fluid from individuals who had undergone SARS-CoV-2 oral fluid molecular RNA testing (Fig. 1A). Participant specimens were collected with informed consent from a diverse population of patients (Fig. 1B). It is important to note that the antibody detection kits presented here do not claim to quantify antibody in the sample but instead aim to provide a binary indicator of antibody presence. Thus, we cannot derive any correlations here between antibody signal and any other factor, but can use the results to determine the presence of an antibody response.

**Figure 1.**
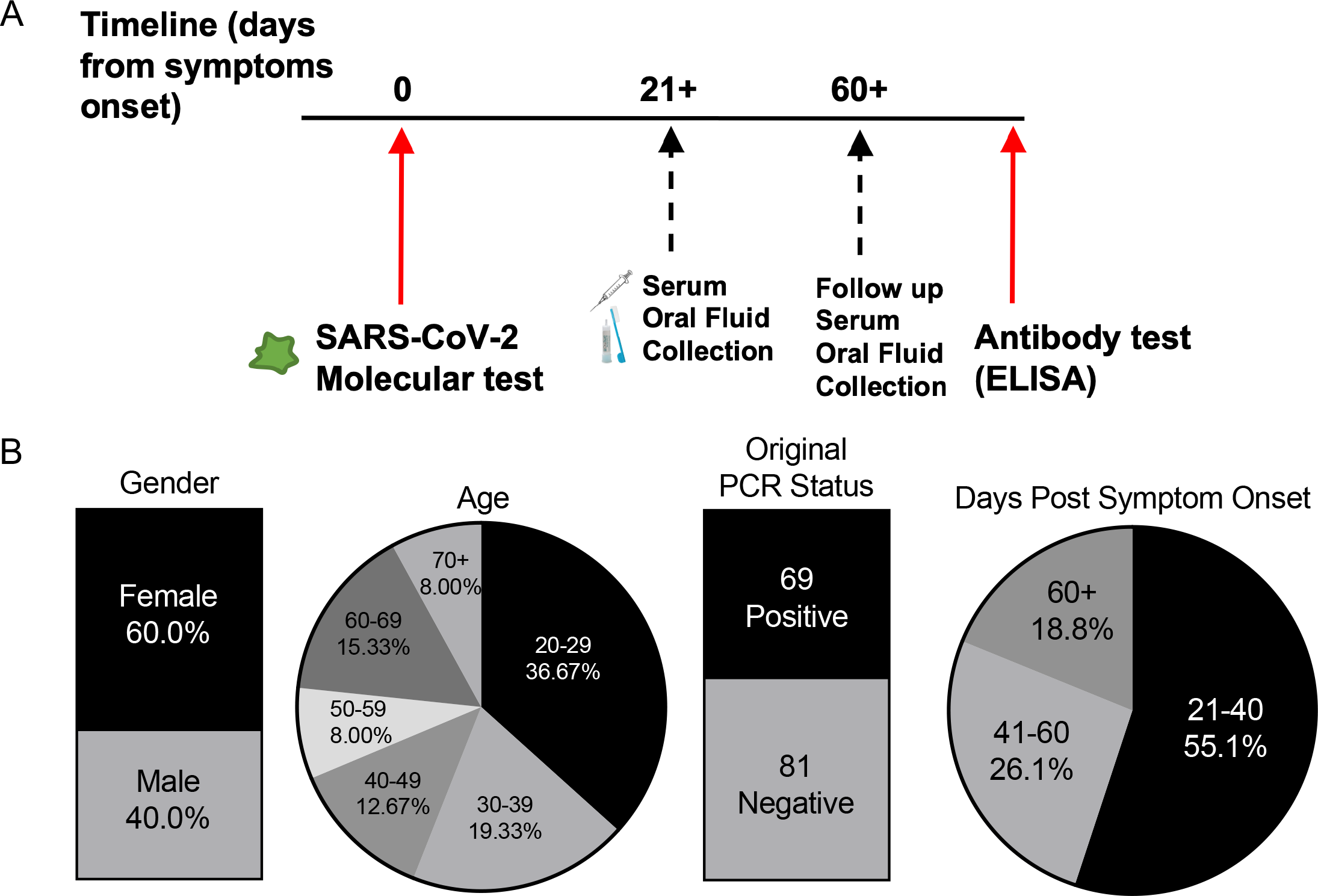
**(A)** Timeline of sample collection, methods and sample metrics from clinical trial participants. **(B)** Visualization of the age, gender, and original PCR status (n = 150) of our cohort of clinical samples, as well as days post symptom onset of our positive participant samples (n = 69).

From positive participants, serum and oral fluid was collected at least 21 days after the date of reported symptom onset to ensure adequate time for antibody seroconversion (2). We tested participant serum specimens on the EuroImmun Anti-SARS-CoV-2 (IgG) ELISA (12) to determine whether the participants were expressing antibodies against the virus using previously described methods (9) (Fig. 2A). Paired oral fluid collected using the OraSure® OACD from sixty-nine SARS-CoV-2 RNA- and serum antibody-positive participants and eighty-one SARS-CoV-2 RNA- and serum antibody-negative participants was evaluated on the OraSure® total antibody ELISA. Antibodies were detected in sixty-four (92.8%) of sixty-nine serum antibody positive participant specimens and zero (100% specificity) of eighty-one serum negative participant specimens (Fig. 2B).

**Figure 2.**
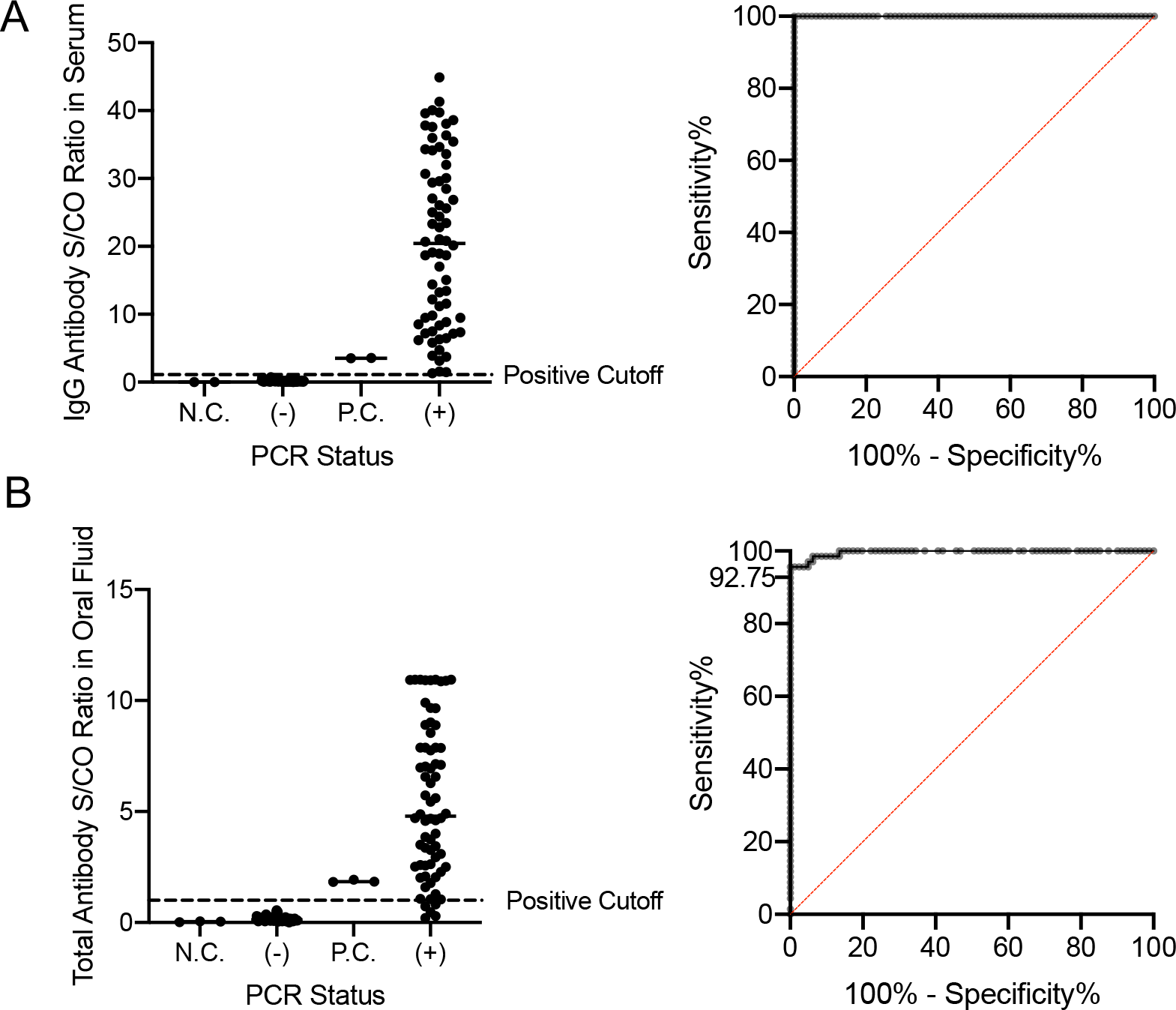
**A)** Detection of antibodies against SARS-CoV-2 in participant-derived serum samples collected from participants who previously tested negative or positive for SARS-CoV-2 RNA by an oral-fluid PCR test. The manufacturer provided positive cutoff value of 1.1 is represented by the dotted line. N.C. = negative control; P.C. = positive control; S/CO = Specimen/Cutoff Ratio. ROC analysis of this dataset in serum results in 100% sensitivity and 100% specificity with an area under the ROC curve (AUC) of 1. **B)** Detection of antibodies against SARS-CoV-2 in paired oral fluid specimens collected using the OraSure® Technologies OACD from the same participants whose serum was tested. The manufacturer provided positive cutoff value of 1.0 is represented by the dotted line. N.C. = negative control; P.C. = positive control; S/CO = Specimen/Cutoff Ratio. ROC analysis of this dataset in oral fluid reveals 92.75% sensitivity and 100% specificity with the manufacturer provided cutoff value of 1.0 total antibody units and an AUC of 0.9965.

We further investigated how well the OACD and ELISA performed when monitoring paired specimens collected from the same participants at later time points post-infection. To evaluate this, we selected sixteen out of our original sixty-nine positive participants and collected their paired oral fluid and serum specimens at least 60 days after the original collection date. We found that the oral fluid-based total antibody ELISA from OraSure® Technologies performed with statistical equivelance in comparison to the serum-based IgG antibody ELISA from EuroImmun, with all positive specimens passing the antibody detection cutoff after both 21+ days and 60+ days (Fig 3.)

**Figure 3.**
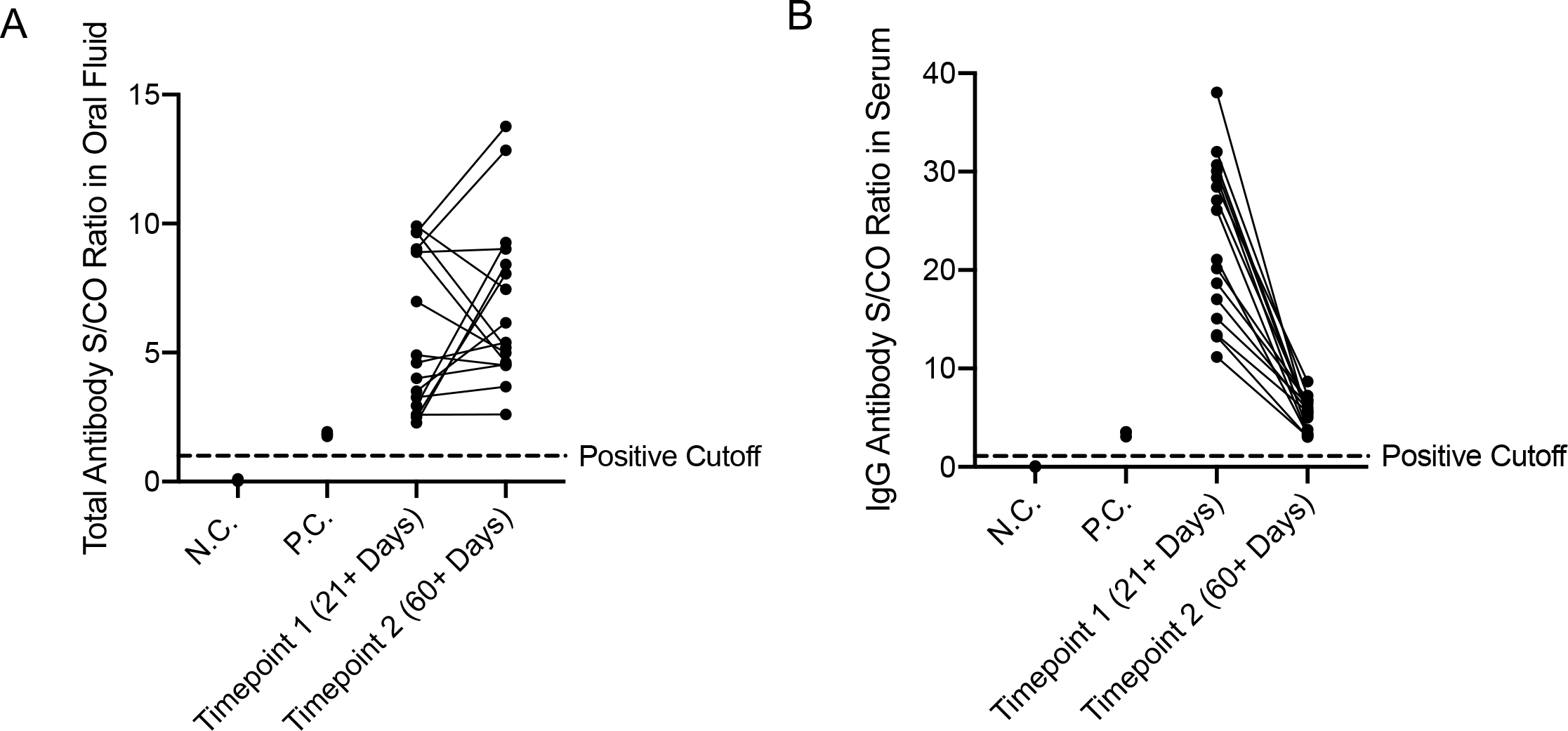
**A)** Detection of antibodies against SARS-CoV-2 in participant-derived serum samples collected from participants who previously tested positive for SARS-CoV-2 RNA by an oral-fluid PCR test both 21+ days post symptom onset and 60+ days post symptom onset (n = 16). All samples passed the positive cutoff value of 1.1 set by the manufacturer and represented by the dotted line. N.C. = negative control; P.C. = positive control; S/CO = Specimen/Cutoff Ratio. ROC analysis of this dataset in serum results in 100% sensitivity and 100% specificity with an area under the ROC curve (AUC) of 1. **B)** Detection of antibodies against SARS-CoV-2 in paired oral fluid samples collected using the OraSure® Technologies OACD from the same participants whose serum was tested (n = 16). The manufacturer provided positive cutoff value of 1.0 is represented by the dotted line. N.C. = negative control; P.C. = positive control; S/CO = Specimen/Cutoff Ratio. ROC analysis of this dataset in oral fluid reveals 100% sensitivity and 100% specificity with the manufacturer provided cutoff value of 1.0 total antibody units and an AUC of 1.

To evaluate the potential time and cost savings of using the OACD for antibody detection instead of traditional serum-based methods, we estimated that a trained phlebotomist operating at a full-capacity drive-thru center can only collect and process blood serum from ∼40 patients in a 5-hour period, resulting in collection of one patient specimen per 7.5 minutes. Because oral fluid can potentially be self-collected using the OACD, face-to-face specimen collection time can be eliminated. This dramatically reduces the amount of time needed for each specimen to be collected, leading us to estimate that we can collect devices from ∼1,000 patients in a similar drive-thru setting over the same 5-hour period, resulting in collection of one specimen in approximately 0.3 minutes. Note that this time does not include the actual time for collection of the sample, which we are assuming individuals would perform while they are waiting in the drive-thru line, thus highlighting the dramatic benefit of self-collected oral samples. This represents approximately a 25-fold increase in sample collection throughput versus traditional blood serum sample collection in the same time frame(Fig. 4).

**Figure 4.**
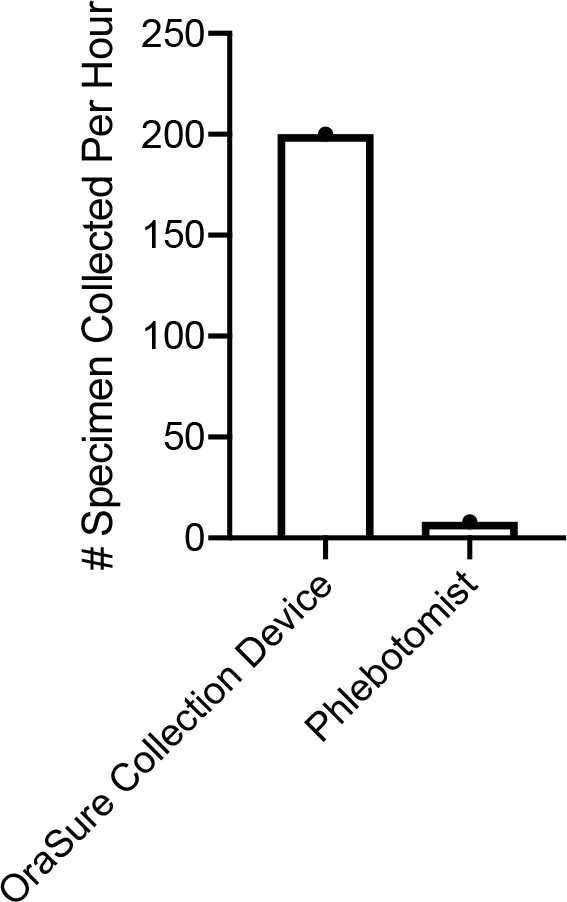
Number of antibody specimens which can be collected in a drive-thru testing center per hour. Oral fluid specimens (200 per hour) are able to be observed self-collected using the OraSure® OACD and simply handed off to a test site employee, while blood serum specimens (8 per hour) must be collected through direct contact with a phlebotomist, thus highlighting the benefit of testing by self-collection.

To increase testing capacity and decrease turnaround time for antibody results, we have automated the process for running the OraSure® Anti-SARS-CoV-2 total antibody ELISA and will continue evaluating the efficiency and reliability of the test.

## Discussion

While antibody testing has previously been limited by the requirement of trained personnel for specimen collection, we have evaluated here a technology for non-invasive self-collection which we believe will be able to be scaled up and mass-produced for widespread testing. Based on the test efficacy and time-savings afforded by this method of antibody detection, OraSure® Technologies has developed a more accessible and affordable testing option for patients interested in understanding whether they have been exposed to and developed antibodies against SARS-CoV-2. The simplicity of the OACD allows us to speculate that this will enable specimens to be theoretically self-collected and eliminate the need for a trained medical associate to administer the collection.

Oral fluid and mucosal antibodies comprise the first line of defense against a respiratory virus and such antibodies are known to typically contribute to sterilizing immunity by neutralizing the virus prior to infection. This neutralization results in both reduced viral load and impact of subsequent infection(13). The results of this study have demonstrated the persistence of antibodies against SARS-CoV-2 in oral fluid of convalescent humans, indicating the importance of having a widely accessible oral fluid antibody detection system to help determine individual immunity in the population.

The OACD, when used with the Anti-SARS-CoV-2 total antibody ELISA kit developed by OraSure® Technologies, has achieved sensitivity and specificity which are equivalent to commercially available serum-based ELISAs(9). This represents a unique and highly successful advancement in antibody detection technology. As we move closer to the development and implementation of a successful vaccine against SARS-CoV-2, we need reliable methods for monitoring antibody production on a wide scale and over time. This antibody detection system allows us to overcome the limitations presented by currently administered serum antibody tests, increasing accessibility to antibody testing and enabling a better understanding of the trends of immune response against this virus over time.

## Methods

### Human serum and oral fluid specimen collection

#### Post-COVID-19 serum and oral fluid specimens

Clinical specimens were collected under the UCLA Institutional Review Board (IRB) approved study protocol IRB#20-000703. The UCLA IRB determined the protocol was of minimal risk and that verbal informed consent was sufficient for the research under 45 CFR 46.117(c). The study team complied with all UCLA policies and procedures, as well as with all applicable Federal, State, and local laws regarding the protection of human subjects in research as stated in the approved IRB. For this study, we obratined three specimens: oral fluid for viral PCR testing, blood via venipuncture, and one oral fluid specimens for antibody testing.

#### Oral fluid swab for viral RNA

This was obtained according to the reported guidelines for Curative’s oral fluid COVID-19 test (14). Briefly, participants coughed hard three times while shielding their cough via mask and/or coughing into the crook of their elbow. They then swabbed the inside of their cheeks, along the top and bottom gums, under the tongue, and finally on the tongue, to gather a sufficient amount of oral fluid. Swabs were placed in a tube containing RNA shield and transported at room temperature before laboratory processing as described. Positive for viral RNA was determined as below 35 cycle threshold (CT).

#### Blood sampling

Participants underwent a standard venipuncture procedure. Briefly, licensed phlebotomists collected a maximum of 15ml whole blood into 3 red-top SST tubes (Becton-Dickinson, cat. number 367988). Once collected, the specimen was left at ambient temperature for 30-60 minutes to coagulate and then was centrifuged at 2200-2500 rpm for 15 minutes at room temperature. Specimens were then placed on ice until delivered to the laboratory site where the serum was aliquoted to appropriate volumes for storage at −80 °C until use.

#### OraSure® Oral Fluid Specimen Collection

OraSure® Oral Antibody Collection Devices (OACD) (item number 3001-3442-70, OraSure®, USA) were used as instructed(10,15). The pad was brushed briefly on the lower and upper gums and then held between the lower gum and the cheek for 2-5 minutes. The pad was then placed into the storage tube, with the provided storage solution. Specimens were kept on ice until they reached the lab. The specimens were processed as recommended by the manufacturer before being aliquoted and stored at −80°C until use(16).

### Serum ELISA

EuroImmun’s FDA-approved SARS-CoV-2 (IgG) ELISA for serum (catalog #EI 2606-9620, EuroImmun, New Jersey, USA) targeting spike (S) protein was run according to the manufacturer provided protocol on the Thunderbolt (Gold Standard Diagnostics, Davis, CA) automated instrument (12). Briefly, serum was diluted 1:101 in each well with the provided sample buffer and then incubated at 37°C for 1 hour. Sample wells were washed three times with a provided wash buffer (10X dilution with ddH2O, 0.35ml per well), before the provided conjugate solution was added (0.1ml per well) and incubated at 37°C for 30 minutes. After a second wash step, the provided substrate solution was added (0.1ml per well) and incubated at ambient temperature for 30 minutes. The provided stop solution was then added (0.1ml per well) and absorbance of sample wells measured immediately at 450 nm and 630 nm, with output reports generated with optical density (O.D.) at 630nm subtracted from O.D. at 450nm.

Data were then analyzed as recommended by the manufacturer and results reported as a ratio (**Equation 1**). Specimens whose ratio exceeded or was equal to 1.1 were considered positive, while specimens with a ratio greater than 0.8 and less than 1.1 were considered equivocal and specimens with a ratio less than or equal to 0.8 were considered negative. Specimens with ratios in the equivocal range were recommended to be rerun, but this did not apply to any specimens in this study.

**Equation 1**. Determination of specimen absorbance ratio based on specimen O.D. divided by the average O.D. of the calibrators.

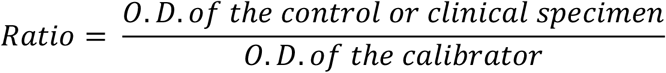

### Oral Fluid ELISA

OraSure® Technologies Anti-SARS-CoV-2 Total Antibody ELISA for oral fluid specimen (item number 3001-3317-70, catalog #1125SB) targeting coronavirus spike protein antigens S1 and S2 was run manually according to the manufacturer provided protocol(11). 25uL of provided sample diluent buffer was added to the desired wells of the plate followed by 100uL of each specimen. Specimens were incubated at ambient temperature for 1 hour. Sample wells were washed six times with a provided wash buffer (20X dilution with ddH2O, 0.35ml per well), before the provided conjugate solution was added (0.1ml per well) and incubated at ambient temperature for 1 hour. After a second wash step, the provided substrate solution was added (0.1ml per well) and incubated at ambient temperature for 30 minutes. The provided stop solution was then added (0.1ml per well) and absorbance of sample wells measured immediately at 450 nm and 630 nm, with output reports generated with O.D. at 630nm subtracted from O.D. at 450nm.

Data were analyzed as recommended by the manufacturer and results reported as a specimen to cutoff (S/CO) ratio (**Equation 2**). Specimens whose ratio was equal to or exceeding 1.0 were considered positive, while specimens with a ratio greater than 0.8 and less than 1.0 were considered equivocal and specimens with a ratio less than or equal to 0.8 were considered negative. Specimens with ratios in the equivocal range were recommended to be rerun, but this did not apply to any specimens in this study.

**Equation 2**. Determination of specimen absorbance ratio based on specimen O.D. divided by the averaged O.D. of the cutoff calibrators.

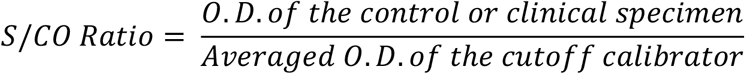

### ELISA Automation

Using the automated Dynex DSX 5 plate ELISA Processing System, we developed a quick two-step automated method for SARS-CoV-2 antibody detection involving: 1) centrifugation of the oral fluid collection devices in a secondary tube to transfer oral fluid in the device and 2) processing of the collected saliva specimens on OraSure® Technologies SARS-CoV-2 Total Antibody ELISA plates.

### Statistical analysis

ROC curves were generated in GraphPad Prism (GraphPad Prism Version 8.4.3, San Diego, USA), with a 95% confidence interval. Area under the curves was also calculated. Correlation between serum and oral fluid specimen IgG values were calculated using a Pearson correlation computing R between the two datasets, with a 95% confidence interval.

### Data availability

The data that support the findings of this study are available from the authors upon reasonable request.

## Data Availability

Full dataset is available upon request

## Acknowledgements

The authors would like to thank Joseph Kapcia III, Cedie Bagos, Aaron Angel, Marilisa Santa-Cruz and Matthew Geluz from Curative, Inc. as well as Kerry Phillips, Toral Zaveri, and Matthew Sullivan from OraSure® Technologies.

## Author Contributions

M.M and designed and ran experiments, analyzed and interpreted data, and drafted the manuscript, P.C. automated the experimental system and drafted the manuscript, A.M. and S.D. analyzed and interpreted data, F.E.T, V.S., and conceptualized the project, A.I. designed experiments, oversaw data collection and analysis, edited the manuscript and maintained correspondence with OraSure® Technologies for the duration of these experiments.

## Figures

**Schematic 1.**
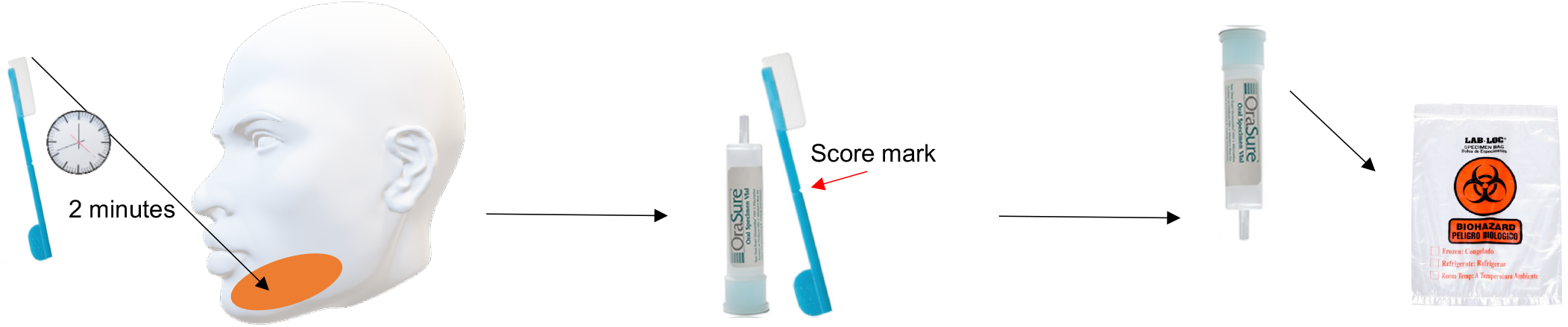
OraSure® Technologies OACD Collection Method. Step 1. Insert collection pad inside the mouth between the lower gum and cheek and gently rub the pad back and forth along the gum line 5 times. Repeat on the other side of the mouth and on both upper gums. Move the pad to the lower gum and leave the pad stationary against the lower gum for 2 minutes. Step 2. Open the collection vial while ensuring enclosed liquid does not spill. Insert collection pad with pad facing down into the liquid in the vial. Bend collection pad handle to break at score mark, discarding the upper half of the stick and place it into the vial. Place the cap on top of the vial and push down firmly until it snaps into place. Step 3. After securing the vial cap, place the vial into a plastic biohazard collection bag and label with patient name and date. This bag will be transported or collected for processing.

